# Bile acid metabolites predict multiple sclerosis progression and supplementation is safe in progressive disease

**DOI:** 10.1101/2024.01.17.24301393

**Authors:** Dimitrios C. Ladakis, Kimystian L. Harrison, Matthew D. Smith, Krista Solem, Sachin Gadani, Larissa Jank, Soonmyung Hwang, Farzaneh Farhadi, Blake E. Dewey, Kathryn C. Fitzgerald, Elias S. Sotirchos, Shiv Saidha, Peter A. Calabresi, Pavan Bhargava

**Affiliations:** Johns Hopkins University School of Medicine, Department of Neurology, Baltimore, United States

## Abstract

**Background:** Bile acid metabolism is altered in multiple sclerosis (MS) and tauroursodeoxycholic acid (TUDCA) supplementation ameliorated disease in mouse models of MS.

**Methods:** Global metabolomics was performed in an observational cohort of people with MS followed by pathway analysis to examine relationships between baseline metabolite levels and subsequent brain and retinal atrophy. A double-blind, placebo-controlled trial, was completed in people with progressive MS (PMS), randomized to receive either TUDCA (2g daily) or placebo for 16 weeks. Participants were followed with serial clinical and laboratory assessments. Primary outcomes were safety and tolerability of TUDCA, and exploratory outcomes included changes in clinical, laboratory and gut microbiome parameters.

**Results:** In the observational cohort, higher primary bile acid levels at baseline predicted slower whole brain, brain substructure and specific retinal layer atrophy. In the clinical trial, 47 participants were included in our analyses (21 in placebo arm, 26 in TUDCA arm). Adverse events did not significantly differ between arms (p=0.77). The TUDCA arm demonstrated increased serum levels of multiple bile acids. No significant differences were noted in clinical or fluid biomarker outcomes. Central memory CD4+ and Th1/17 cells decreased, while CD4+ naïve cells increased in the TUDCA arm compared to placebo. Changes in the composition and function of gut microbiota were also noted in the TUDCA arm compared to placebo.

**Conclusion:** Bile acid metabolism in MS is linked with brain and retinal atrophy. TUDCA supplementation in PMS is safe, tolerable and has measurable biological effects that warrant further evaluation in larger trials with a longer treatment duration.

**Trial registration:** ClinicalTrials.gov NCT03423121

**Funding:** National MS Society grant RG-1707-28601 to PB, R01 NS082347 from NINDS to PAC and National MS Society grant RG-1606-08768 to SS.

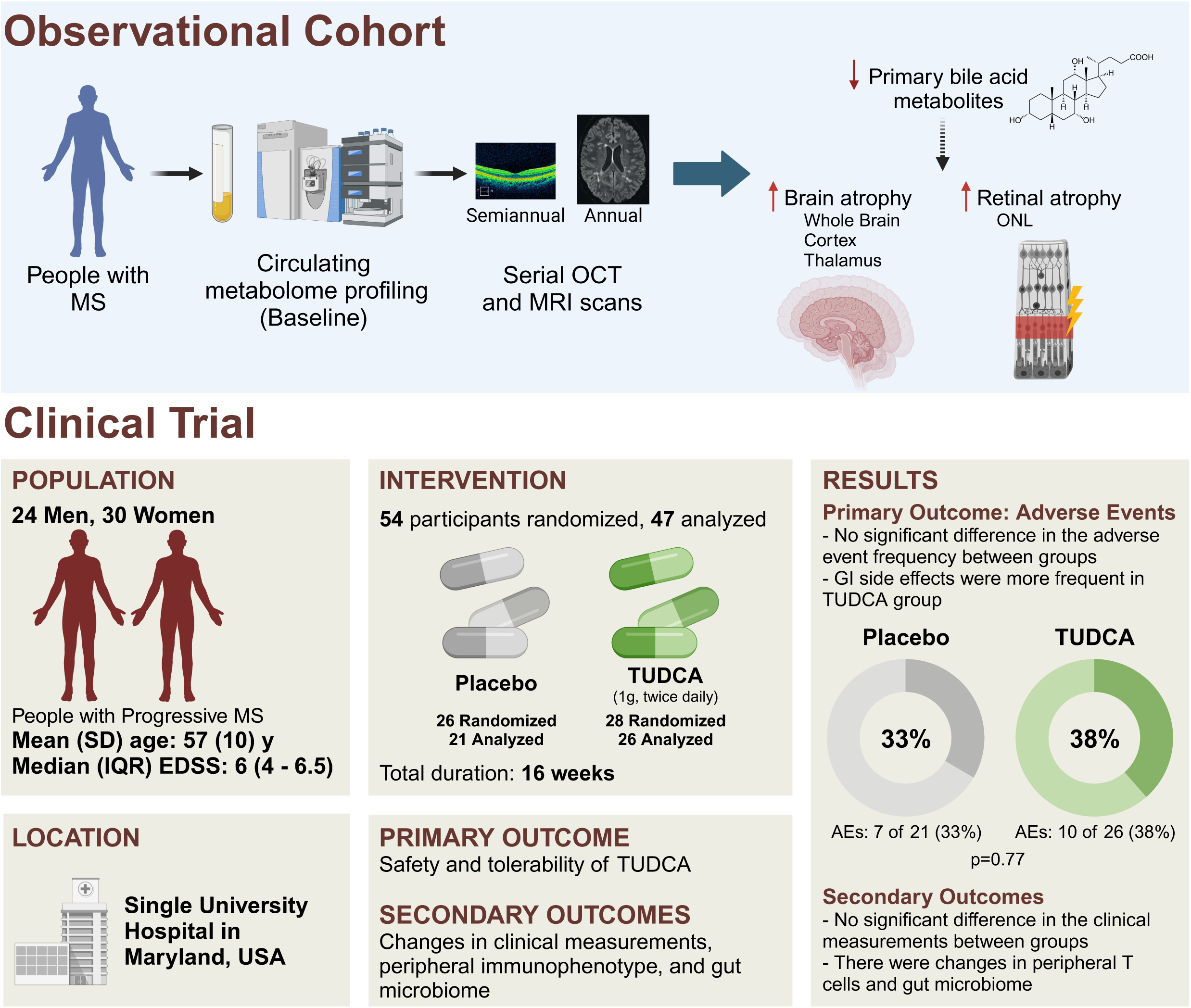

## INTRODUCTION

Multiple sclerosis (MS) is an inflammatory disorder of the central nervous system (CNS) that results in demyelination and neurodegeneration (1). Altered levels of circulating bile acid metabolites were identified in a cross-sectional study involving both adult and pediatric-onset MS populations (2–5). People with progressive MS (PMS) were found to have more notable reductions in bile acid levels compared to relapsing remitting MS (RRMS) (2). Whether levels of circulating bile acid metabolites predict subsequent MS disease course is unknown.

In an animal model of MS, supplementation with a secondary bile acid, tauroursodeoxycholic acid (TUDCA), resulted in reduced inflammation and ameliorated disease severity (2). We also noted direct anti-inflammatory effects of TUDCA on astrocytes and microglia *in vitro*. Similar results and neuroprotective effects have been identified with bile acid supplementation in models of other neurodegenerative conditions, however the mechanism of action is unclear (6–9). Bile acid receptors such as GPBAR1, may be a potential target, as these are present in the CNS on the surface of astrocytes and myeloid cells within white matter MS lesions (2). Clinical trials in other neurodegenerative diseases have shown that bile acid supplementation is safe, well-tolerated, can lead to increased levels of bile acids in the CNS, and has beneficial effects on clinical outcomes leading to FDA approval of a bile acid compound for amyotrophic lateral sclerosis (10–14). However, the effects of bile acid supplementation in people with MS are unknown.

In this manuscript we first describe the relationship between baseline circulating bile acid metabolite levels and subsequent CNS atrophy based on imaging outcomes (MRI and optical coherence tomography [OCT]). We then present the results of a phase 1/2a placebo-controlled randomized double-blind trial of TUDCA supplementation in people with PMS (2).

## RESULTS

### Circulating bile acid levels predict subsequent retinal and brain atrophy

We identified people with MS with baseline circulating bile acid level data followed by serial MRI (N=107) and/or serial optical coherence tomography (OCT) scans (N=192). Demographics for this cohort can be found in Table 1. Using linear mixed effects models adjusted for age at baseline, sex and MS subtype, followed by pathway enrichment analysis, we noted that higher baseline primary bile acid (synthesized in the liver from cholesterol) metabolite levels were associated with slower cerebral white matter (normalized enrichment score (NES)=1.6; p=0.047), thalamic (NES=2; p=5E-04), pallidum (NES=1.8; p=0.01), subcortical gray matter (NES=1.7; p=0.01), and whole brain (NES=1.7; p=0.01) atrophy. In similar analyses, higher levels of secondary bile acid metabolites were associated with faster T2 lesion burden accumulation over time (NES=2.2; p=3E-04). In the OCT cohort using the same analytical methods, people with higher primary bile acid metabolite levels demonstrated slower outer nuclear layer (ONL) atrophy (NES=1.7; p=0.002). No association was found with ganglion cell and inner plexiform (GCIPL) or inner nuclear (INL) layers (Table 2). Individual metabolite results can be found in Supplemental Tables 1 (MRI results) and 2 (OCT results). Overall, these data demonstrate that baseline levels of circulating bile acid metabolites are associated with subsequent MS progression based on imaging metrics.

**Table 1.** Demographics and clinical characteristics of the observational cohort.

| **Characteristic** | **MRI cohort**<br>**N = 107<sup>I</sup>** | **OCT cohort**<br>**N = 192<sup>I</sup>** |
| --- | --- | --- |
| **Sex, Male, n (%)** | 29 (27%) | 51 (27%) |
| **Age, years, mean (SD)** | 43.2 (12.9) | 43.3 (12.2) |
| **Race, n (%)** |  |  |
| Black | 11 (10%) | 19 (10%) |
| White | 91 (85%) | 167 (87%) |
| Other | 5 (5%) | 5 (3%) |
| **MS subtype, n (%)** |  |  |
| PMS | 30 (28%) | 53 (27%) |
| RRMS | 77 (72%) | 139 (73%) |
| **Follow up, years, median (IQR)** | 5.3 (3.2 to 7.8) | 5.6 (3.9 to 8) |
| **Eyes with history of ON, n (%)** | - | 118 (33%) |

**Table 2.**
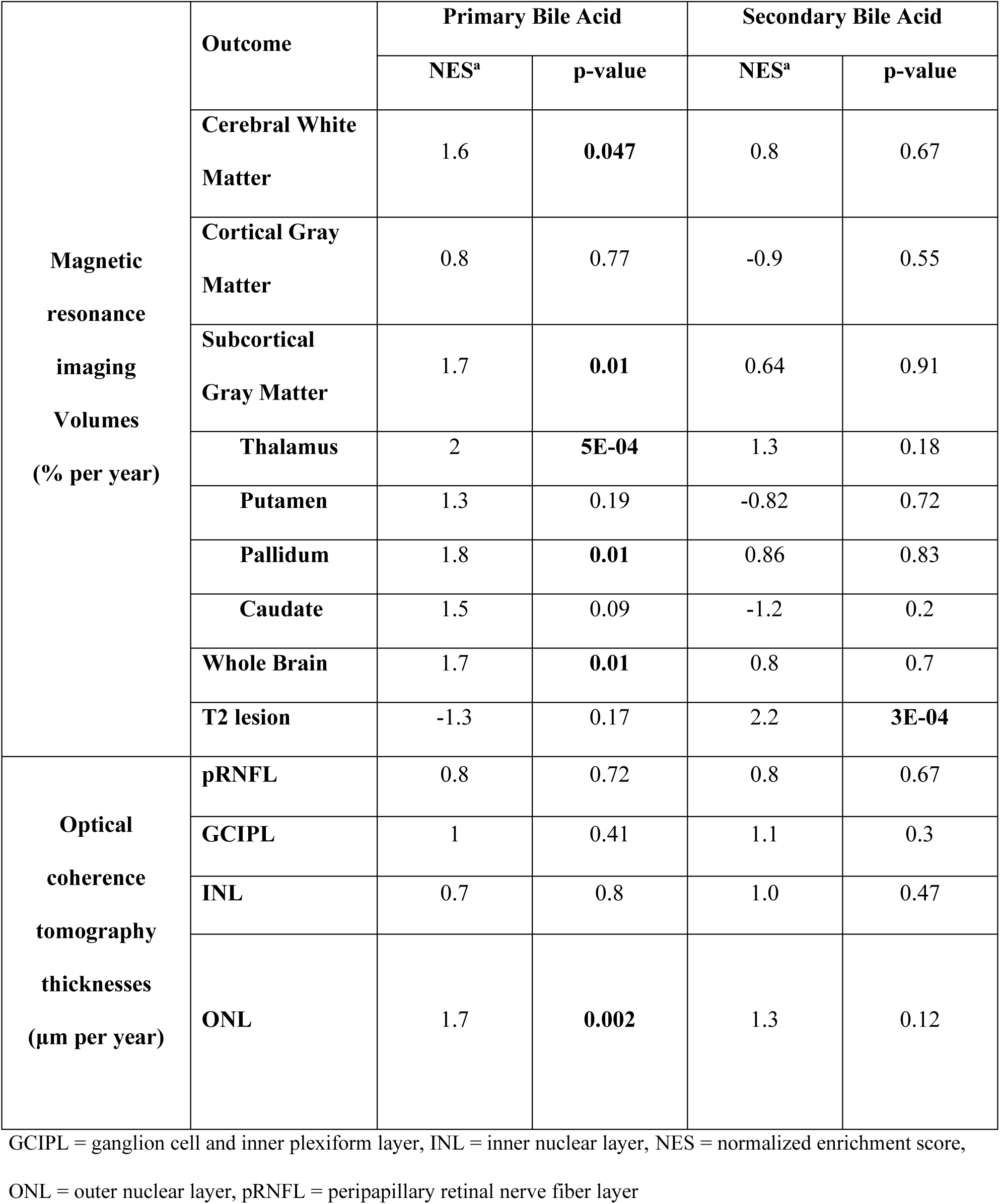
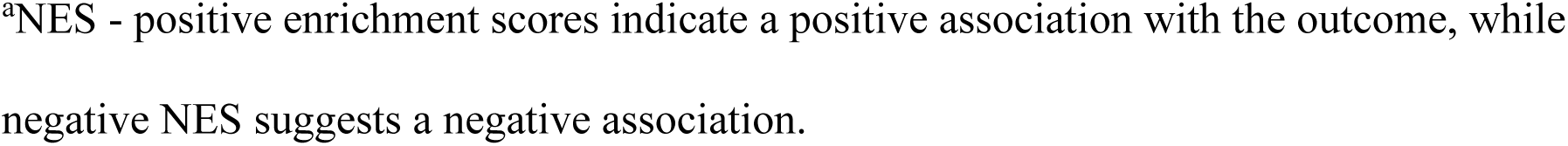
Associations of bile acid metabolite pathways and imaging outcomes.

|  | Outcome | Primary Bile Acid |  | Secondary Bile Acid |  |
| --- | --- | --- | --- | --- | --- |
|  |  | NES<sup>a</sup> | p-value | NES<sup>a</sup> | p-value |
| **Magnetic resonance imaging Volumes (% per year)** | **Cerebral White Matter** | 1.6 | **0.047** | 0.8 | 0.67 |
|  | **Cortical Gray Matter** | 0.8 | 0.77 | -0.9 | 0.55 |
|  | **Subcortical Gray Matter** | 1.7 | **0.01** | 0.64 | 0.91 |
|  | **Thalamus** | 2 | **5E-04** | 1.3 | 0.18 |
|  | **Putamen** | 1.3 | 0.19 | -0.82 | 0.72 |
|  | **Pallidum** | 1.8 | **0.01** | 0.86 | 0.83 |
|  | **Caudate** | 1.5 | 0.09 | -1.2 | 0.2 |
|  | **Whole Brain** | 1.7 | **0.01** | 0.8 | 0.7 |
|  | **T2 lesion** | -1.3 | 0.17 | 2.2 | **3E-04** |
| **Optical coherence tomography thicknesses (µm per year)** | **pRNFL** | 0.8 | 0.72 | 0.8 | 0.67 |
|  | **GCIPL** | 1 | 0.41 | 1.1 | 0.3 |
|  | **INL** | 0.7 | 0.8 | 1.0 | 0.47 |
|  | **ONL** | 1.7 | **0.002** | 1.3 | 0.12 |

GCIPL = ganglion cell and inner plexiform layer, INL = inner nuclear layer, NES = normalized enrichment score,
ONL = outer nuclear layer, pRNFL = peripapillary retinal nerve fiber layer
<sup>a</sup>NES - positive enrichment scores indicate a positive association with the outcome, while negative NES suggests a negative association.

### Study population of the randomized double-blind placebo-controlled trial of TUDCA supplementation

Between July 2018 and April 2022, 59 patients were screened for the study, of whom 54 met the eligibility criteria, were enrolled, and randomized into the study – 26 to the placebo arm and 28 to the TUDCA arm. Of the 54 participants enrolled, 47 (87%) completed two or more visits, of whom 21 were in the placebo arm and 26 in the TUDCA arm (Figure 1). These people were specified as meeting eligibility criteria for analytical models and a modified intention-to-treat approach was used for analyses. Baseline demographics and clinical characteristics of the two arms are shown in Table 3. Overall, the study cohort consisted of 30 (56%) females and 24 (44%) males, with a mean age of 57 years. All the participants were white and there were 26 (48%) people with primary PMS and 28 (52%) with secondary PMS. About half the people (26) were on infused disease-modifying therapies (DMT) and 25 were not on DMT. The remainder were on either injectable (two) or oral (one) DMTs.

**Figure 1.**
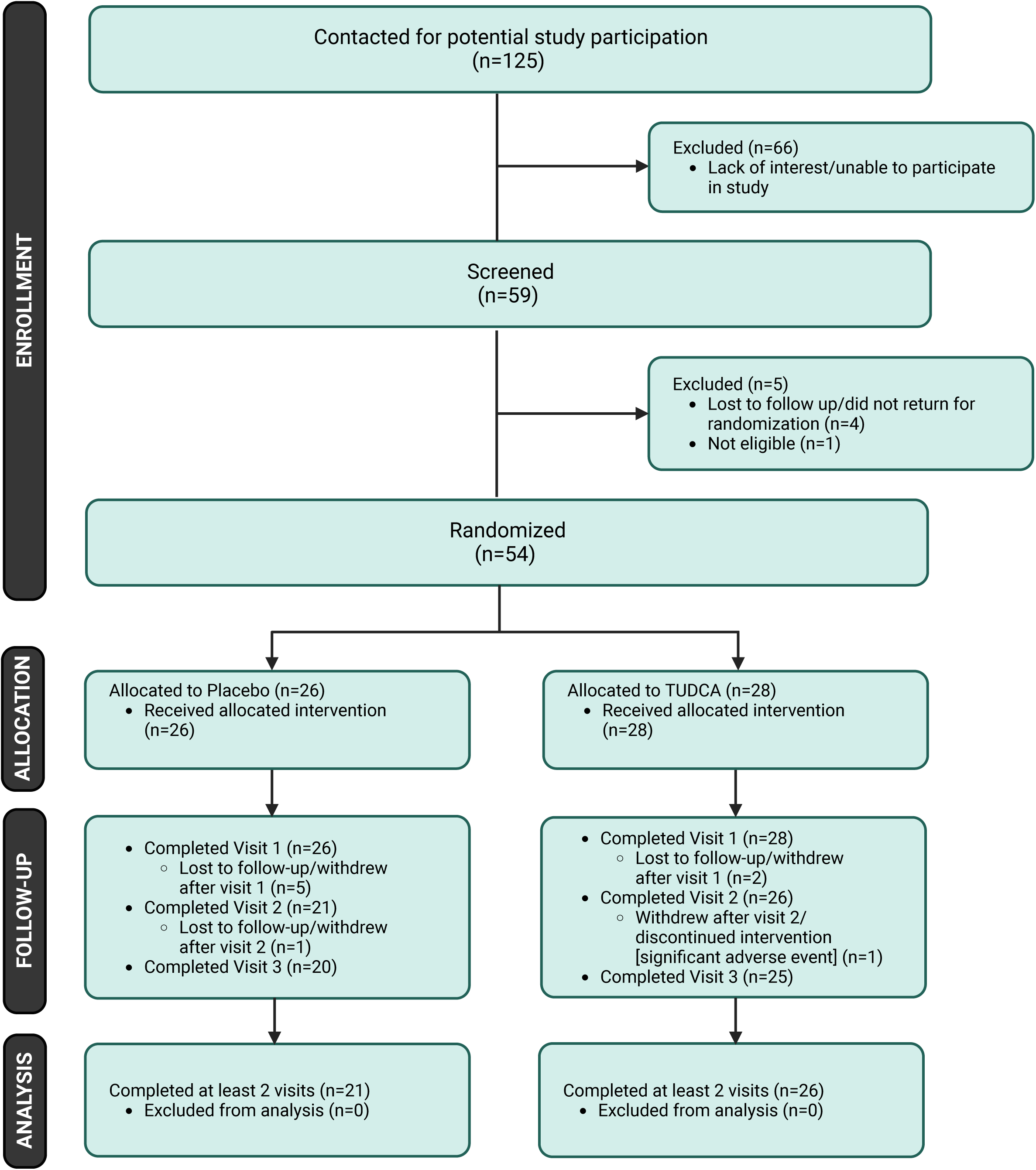
Patient Flow Chart for the Clinical Trial. Screening, randomization and follow-up of the participants. Five participants in the placebo arm and two participants in the TUDCA arm did not have a follow-up visit and were therefore excluded from analyses. Created with BioRender.com.

**Table 3.** Demographics and clinical characteristics of clinical trial cohort.

| **Characteristic** | **Placebo, N = 26** | **TUDCA, N = 28** |
| --- | --- | --- |
| **Female, n (%)** | 16 (62%) | 14 (50%) |
| **Age, Median (IQR)** | 58.3 (10.0) | 55.7 (10.8) |
| **MS Subtype, n (%)** |  |  |
| Primary Progressive | 10 (38%) | 16 (57%) |
| Secondary Progressive | 16 (62%) | 12 (43%) |
| **DMT, n (%)** |  |  |
| Dimethyl Fumarate | 1 (3.8%) | 0 (0%) |
| Natalizumab | 2 (7.7%) | 2 (7.1%) |
| Glatiramer Acetate | 1 (3.8%) | 1 (3.6%) |
| Ocrelizumab | 9 (35%) | 9 (32%) |
| Rituximab | 1 (3.8%) | 3 (11%) |
| None | 12 (46%) | 13 (46%) |
| **EDSS, Median (IQR)<sup>a</sup>** | 6.0 (5.1 to 6.5) | 6.0 (3.9 to 6.5) |
| **Symptoms Duration,**<br>**Median (IQR)** | 22 (14 to 29) | 13 (6 to 24) |
| **Disease Duration, Median**<br>**(IQR)** | 15 (9 to 23) | 8 (2 to 14) |

### TUDCA supplementation was safe and tolerable in PMS

Adverse events (AEs) and their incidence can be found in Table 4. AEs occurred comparably in the two groups, with 7 (33%) participants in the placebo arm experiencing a total of 8 AEs and 10 (38%) participants in the TUDCA arm reporting 11 AEs. Gastrointestinal (GI)-related AEs (including nausea/vomiting, diarrhea, indigestion/reflux, abdominal pain/cramps, and cholangitis) were the majority of the AEs in the TUDCA group. There were 4 (19%) GI-related adverse events in the placebo group (including indigestion/reflux, constipation, abdominal pain/cramps, and increased flatulence). Notably, the most common side effect in the TUDCA group was diarrhea, which occurred in three participants while no one in the placebo group developed this adverse event. One non-GI-related AE (facial rash) occurred in the TUDCA arm, compared to 4 in the placebo arm (including urinary tract infection, upper respiratory infection, and deep venous thrombosis). There was one serious AE (cholangitis) reported in the TUDCA group, which also led to hospitalization; it resolved without sequelae and was deemed to be unlikely associated to the study treatment by the data and safety monitoring board (DSMB) of the study, as the patient was found to have magnetic resonance cholangiopancreatography (MRCP) showing chronic changes including abnormalities in the biliary tree and evidence of fibrosis/cirrhosis in addition to a hepatobiliary iminodiacetic acid (HIDA) scan that showed no emptying of the gall bladder which was felt to represent chronic cholecystitis. There were no serious AEs or AEs leading to hospitalization in the placebo group. Thus, we demonstrate that TUDCA supplementation at a dose of 1000 mg twice daily for a duration of four months is safe and tolerable in people with PMS.

**Table 4.** Adverse Events.

|  | **Placebo**<br>N = 21 | **TUDCA**<br>N = 26 | **p-value** |
| --- | --- | --- | --- |
| **Distinct adverse events, n (%)** | 8 (38%) | 11 (42%) | - |
| **Patients with $\geq 1$ adverse event, n (%)** | 7 (33%) | 10 (38%) | 0.77 |
| **GI-related adverse event, n (%)** | 4 (19%) | 10 (38%) | 0.2 |
| **Nausea/Vomiting** | - | 2 (8%) |  |
| **Diarrhea** | - | 3 (12%) |  |
| **Indigestion/Reflux** | 1 (5%) | 2 (8%) |  |
| **Constipation** | 1 (5%) | - |  |
| **Abdominal pain/cramps** | 1 (5%) | 2 (8%) |  |
| **Increased flatulence** | 1 (5%) | - |  |
| **Cholangitis** | - | 1 (4%) |  |
| **Non-GI adverse event, n (%)** | 4 (19%) | 1 (3.8%) | 0.16 |
| **Urinary tract infection** | 2 (10%) | - |  |
| **Upper respiratory infection** | 1 (5%) | - |  |
| **Deep venous thrombosis** | 1 (5%) | - |  |
| **Facial Rash** | - | 1 |  |
| **Adverse event leading to hospitalization, n (%)** | 0 (0%) | 1 (3.8%) | >0.99 |
| **Any serious adverse event, n (%)** | 0 (0%) | 1 (3.8%) | >0.99 |

<sup>a</sup>P-values derived from Fisher's exact test

### TUDCA supplementation altered circulating bile acid levels in PMS

Targeted metabolomics analyses were performed using serum obtained at each visit to determine the effects of treatment on the circulating metabolome. The metabolomics panel included 15 primary and secondary bile acids. Table 5 demonstrates the rates of change in bile acids over time in each group, as well as the between-group difference as derived from linear mixed-effects models. The TUDCA arm demonstrated significant increases in five out of the 15 bile acid levels tested over the study period, including levels of TUDCA (log(change) per 16 weeks beta=2.52, 95% CI: 1.3 to 3.73; p=1.51E-04), glycoursodeoxycholic acid (GUDCA; beta=2.3, 95% CI: 1.32 to 3.29; p=2.78E-05), ursodeoxycholic acid (UDCA; beta=1.89, 95% CI: 0.76 to 3.02; p=0.001), lithocholic acid (LCA; beta=0.75, 95% CI: 0.28 to 1.22; p=0.002) and glycolithocholic acid (GLCA; beta=0.9, 95% CI: 0.33 to 1.47; p=0.003). The rates of change in LCA and GLCA were not statistically different from those in the placebo group. These data demonstrate that TUDCA supplementation results in increase of multiple circulating bile acid levels in PMS.

**Table 5.**
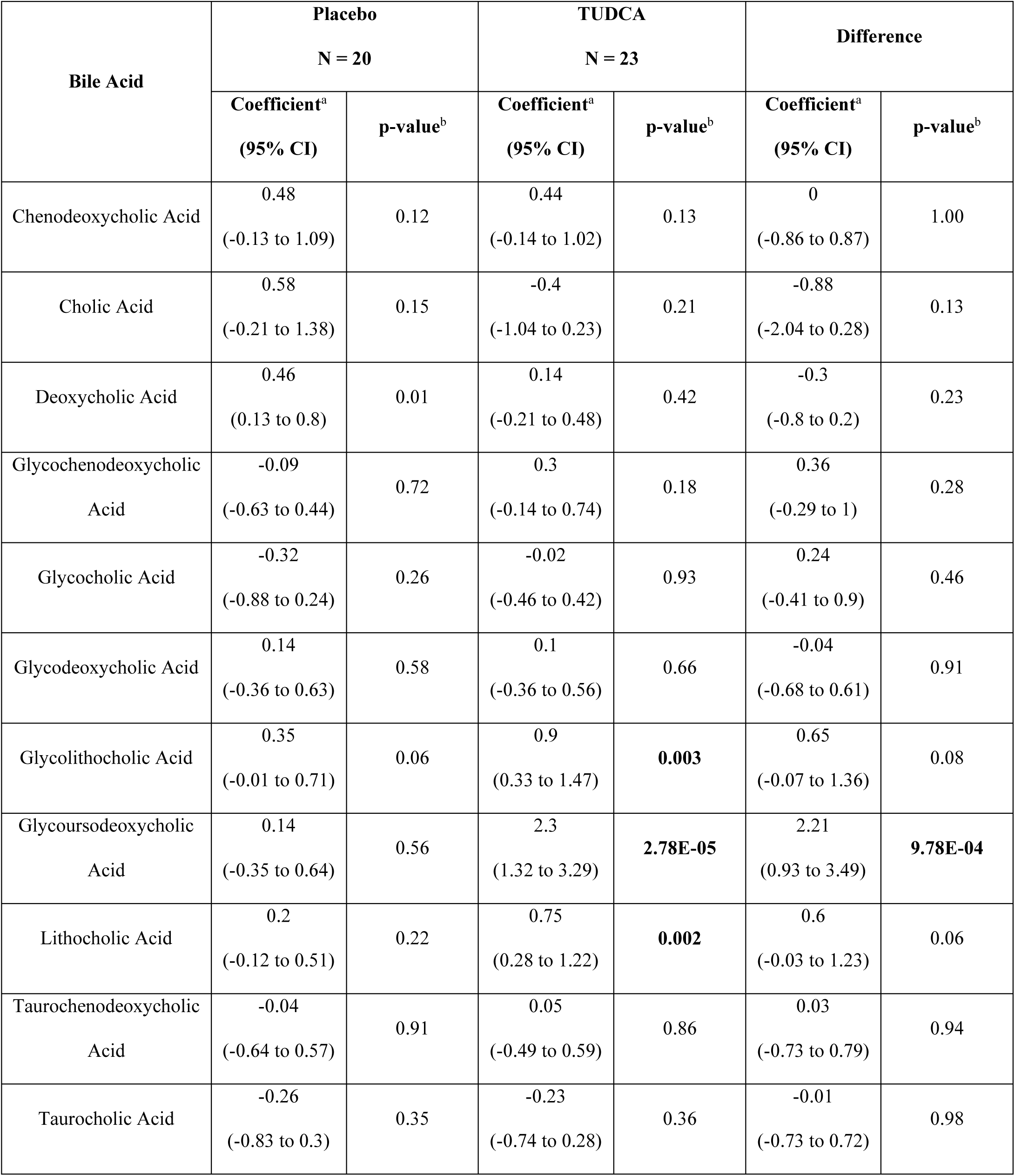
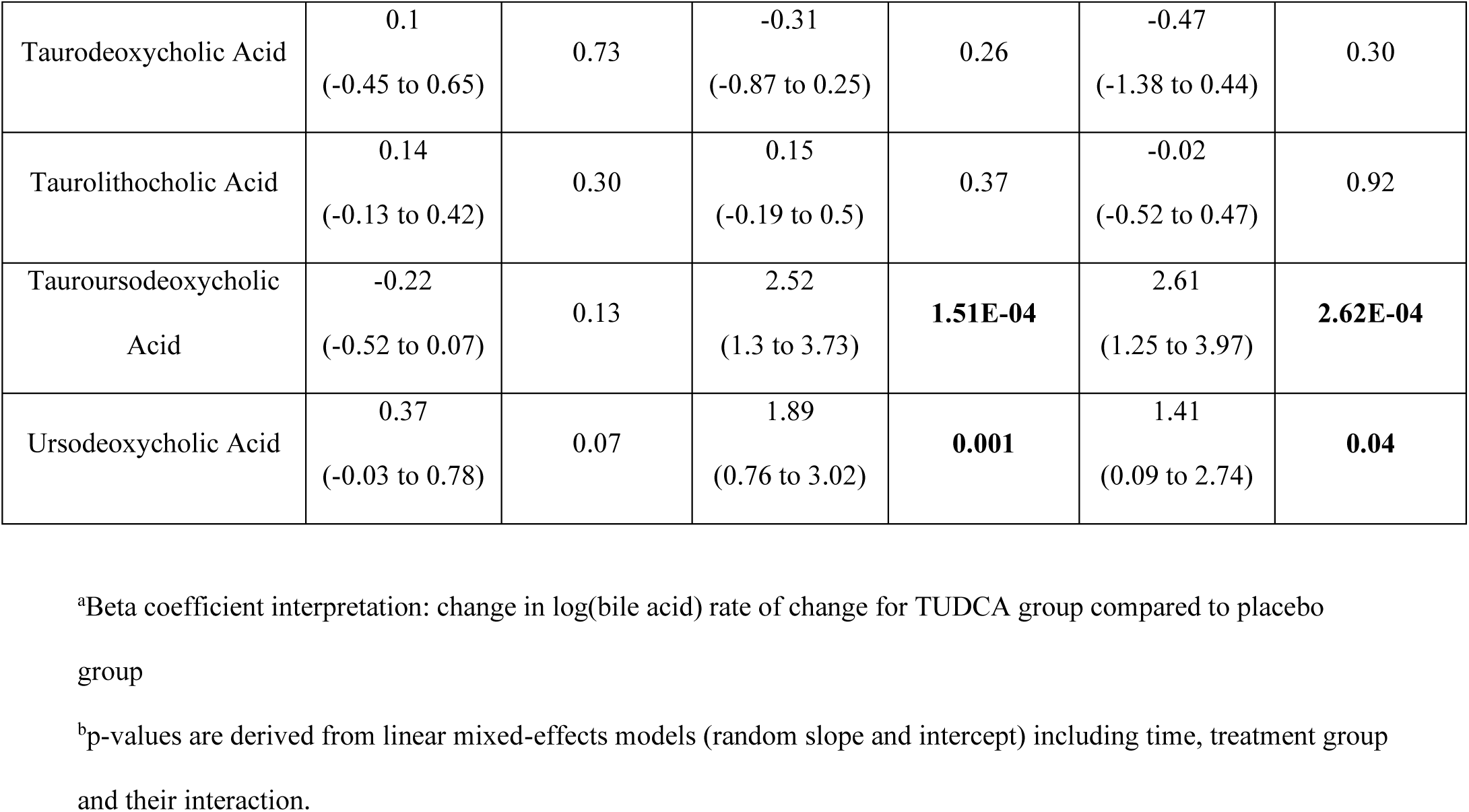
Changes in bile acid levels over time (change per 16 weeks)

| Bile Acid | Placebo<br>N = 20 |  | TUDCA<br>N = 23 |  | Difference |  |
| --- | --- | --- | --- | --- | --- | --- |
|  | Coefficient<sup>a</sup><br>(95% CI) | p-value<sup>b</sup> | Coefficient<sup>a</sup><br>(95% CI) | p-value<sup>b</sup> | Coefficient<sup>a</sup><br>(95% CI) | p-value<sup>b</sup> |
| Chenodeoxycholic Acid | 0.48<br>(-0.13 to 1.09) | 0.12 | 0.44<br>(-0.14 to 1.02) | 0.13 | 0<br>(-0.86 to 0.87) | 1.00 |
| Cholic Acid | 0.58<br>(-0.21 to 1.38) | 0.15 | -0.4<br>(-1.04 to 0.23) | 0.21 | -0.88<br>(-2.04 to 0.28) | 0.13 |
| Deoxycholic Acid | 0.46<br>(0.13 to 0.8) | 0.01 | 0.14<br>(-0.21 to 0.48) | 0.42 | -0.3<br>(-0.8 to 0.2) | 0.23 |
| Glycochenodeoxycholic Acid | -0.09<br>(-0.63 to 0.44) | 0.72 | 0.3<br>(-0.14 to 0.74) | 0.18 | 0.36<br>(-0.29 to 1) | 0.28 |
| Glycocholic Acid | -0.32<br>(-0.88 to 0.24) | 0.26 | -0.02<br>(-0.46 to 0.42) | 0.93 | 0.24<br>(-0.41 to 0.9) | 0.46 |
| Glycodeoxycholic Acid | 0.14<br>(-0.36 to 0.63) | 0.58 | 0.1<br>(-0.36 to 0.56) | 0.66 | -0.04<br>(-0.68 to 0.61) | 0.91 |
| Glycolithocholic Acid | 0.35<br>(-0.01 to 0.71) | 0.06 | 0.9<br>(0.33 to 1.47) | **0.003** | 0.65<br>(-0.07 to 1.36) | 0.08 |
| Glycoursodeoxycholic Acid | 0.14<br>(-0.35 to 0.64) | 0.56 | 2.3<br>(1.32 to 3.29) | **2.78E-05** | 2.21<br>(0.93 to 3.49) | **9.78E-04** |
| Lithocholic Acid | 0.2<br>(-0.12 to 0.51) | 0.22 | 0.75<br>(0.28 to 1.22) | **0.002** | 0.6<br>(-0.03 to 1.23) | 0.06 |
| Taurochenodeoxycholic Acid | -0.04<br>(-0.64 to 0.57) | 0.91 | 0.05<br>(-0.49 to 0.59) | 0.86 | 0.03<br>(-0.73 to 0.79) | 0.94 |
| Taurocholic Acid | -0.26<br>(-0.83 to 0.3) | 0.35 | -0.23<br>(-0.74 to 0.28) | 0.36 | -0.01<br>(-0.73 to 0.72) | 0.98 |
| Taurodeoxycholic Acid | 0.1<br>(-0.45 to 0.65) | 0.73 | -0.31<br>(-0.87 to 0.25) | 0.26 | -0.47<br>(-1.38 to 0.44) | 0.30 |
| Taurolithocholic Acid | 0.14<br>(-0.13 to 0.42) | 0.30 | 0.15<br>(-0.19 to 0.5) | 0.37 | -0.02<br>(-0.52 to 0.47) | 0.92 |
| Tauroursodeoxycholic<br>Acid | -0.22<br>(-0.52 to 0.07) | 0.13 | 2.52<br>(1.3 to 3.73) | **1.51E-04** | 2.61<br>(1.25 to 3.97) | **2.62E-04** |
| Ursodeoxycholic Acid | 0.37<br>(-0.03 to 0.78) | 0.07 | 1.89<br>(0.76 to 3.02) | **0.001** | 1.41<br>(0.09 to 2.74) | **0.04** |

<sup>a</sup>Beta coefficient interpretation: change in log(bile acid) rate of change for TUDCA group compared to placebo group
<sup>b</sup>p-values are derived from linear mixed-effects models (random slope and intercept) including time, treatment group and their interaction.

### Effects of TUDCA supplementation on clinical outcomes and soluble biomarkers

Table 6 displays the changes in clinical outcome measures over time between the two groups. Although there was a significant improvement in overall multiple sclerosis functional composite (MSFC) scores (beta=0.14, 95%CI: 0.02 to 0.27; p=0.03) over the study period in the TUDCA group, there was no significant difference in the rates of change in the MSFC score between the TUDCA and placebo group (beta=0.11, 95%CI: -0.07 to 0.3; p=0.22). There were no significant changes observed in the EDSS and the MSQOL-54 (physical and mental components) in either group, or in the rate of change of these clinical outcomes between the two groups (Table 6).

**Table 6.** Comparison of Clinical Outcomes and Biomarker Changes (change per 16 weeks)

| Clinical outcome | Placebo |  |  | TUDCA |  |  | Difference |  |  |
| --- | --- | --- | --- | --- | --- | --- | --- | --- | --- |
|  | N | Coefficient<sup>a</sup><br>(95% CI) | p-value<sup>b</sup> | N | Coefficient<sup>a</sup><br>(95% CI) | p-value<sup>b</sup> | N | Coefficient<sup>a</sup><br>(95% CI) | p-value<sup>c</sup> |
| EDSS | 21 | -0.22 (-0.58 to 0.13) | 0.21 | 23 | 0.19 (-0.11 to 0.49) | 0.20 | 44 | 0.34 (-0.08 to 0.75) | 0.11 |
| Timed 25-foot walk test z-score | 20 | 0 (-0.03 to 0.03) | 0.92 | 23 | 0.01 (-0.02 to 0.03) | 0.62 | 43 | 0 (-0.03 to 0.03) | 1.00 |
| 9-hole peg test z-score | 21 | -0.05 (-0.16 to 0.06) | 0.34 | 23 | 0.07 (-0.05 to 0.2) | 0.25 | 44 | 0.12 (-0.04 to 0.29) | 0.14 |
| PASAT z-score | 18 | 0.16 (-0.16 to 0.48) | 0.31 | 22 | 0.37 (0 to 0.73) | 0.05 | 40 | 0.25 (-0.26 to 0.77) | 0.33 |
| MSFC z-score | 17 | 0.04 (-0.08 to 0.17) | 0.51 | 22 | 0.14 (0.02 to 0.27) | **0.03** | 39 | 0.11 (-0.07 to 0.3) | 0.22 |
| MSQOL – Physical | 17 | 0.06 (-4.65 to 4.78) | 0.98 | 18 | 1.04 (-3.96 to 6.03) | 0.67 | 35 | 0.91 (-5.44 to 7.25) | 0.78 |
| MSQOL - Mental | 19 | 4.6 (-0.16 to 9.35) | 0.06 | 19 | -0.1 (-4.69 to 4.48) | 0.96 | 38 | -4.89 (-11.28 to 1.49) | 0.13 |
| GFAP (log change) | 20 | 0.12 (-0.12 to 0.36) | 0.32 | 23 | 0.02 (-0.17 to 0.2) | 0.87 | 43 | -0.12 (-0.36 to 0.12) | 0.31 |
| NfL (log change) | 20 | 0.07 (-0.17 to 0.31) | 0.56 | 23 | 0.05 (-0.16 to 0.26) | 0.65 | 43 | -0.07 (-0.34 to 0.2) | 0.62 |

<sup>a</sup> Beta coefficient interpretation: change in dependent variable per 16 weeks for TUDCA group compared to placebo group
<sup>b</sup> P-values derived from linear mixed-effects models (random slope) including time
<sup>c</sup> P-values derived from linear mixed-effects models (random slope) including treatment group with time interaction

TUDCA administration did not have an effect on rates of change of serum neurofilament light-chain (NfL) (difference in log(change): -0.07, 95% CI: -0.34 to 0.2; p=0.62) or glial fibrillary acidic protein (GFAP) (difference in log(change): -0.12, 95% CI: -0.36 to 0.12; p=0.31) levels.

Overall, we did not note significant changes in clinical or biomarker outcomes with TUDCA supplementation compared to placebo over the course of the trial which was not unexpected given the trial duration.

### TUDCA supplementation impacts the peripheral immunophenotype

Changes in the proportion of peripheral immune cell populations (assessed using multiparametric flowcytometry) over time between the two groups are shown in Table 7. Participants in the TUDCA group demonstrated a decrease in central memory CD4+ T cells (CD3+;CD56- ;CD4+CD45RA-;CCR7+, difference in rate of change compared to placebo: -4.9%, 95% CI: -7.9% to -1.9%, p=0.002), Th1/17 (double-positive) cells (CD3+;CD56-;CD4+;CCR4-;CXCR3+,CCR6+, difference in rate of change compared to placebo: -2.8%, 95% CI: -4.9% to -0.6%, p=0.01), Th1 cells (CD3+;CD56-;CD4+;CCR4-,CXCR3+;CCR6-, difference in rate of change compared to placebo: -2.6%, 95% CI: -5% to -0.2%, p=0.03) and regulatory T (Treg) cells (CD3+;CD56-;CD4+;FoxP3+;CD25hi, difference in rate of change compared to placebo: -0.6%, 95% CI: -1.1% to -0.03%, p=0.04) over the study period. On the other hand, there was an increase in CD4+ naïve cells (CD3+;CD56-;CD4+;CD45RA+;CCR7+, difference in rate of change compared to placebo: 6.5%, 95% CI: 1.9% to 11.1%, p=0.01) in the TUDCA arm compared to placebo arm (Figure 2). There was no correlation between the rate of change in TUDCA levels and the rate of change in immune cell subsets. Thus, we found that TUDCA supplementation had significant impacts on T cell populations specifically reducing memory T cells and other inflammatory T cell sub-types.

**Figure 2.**
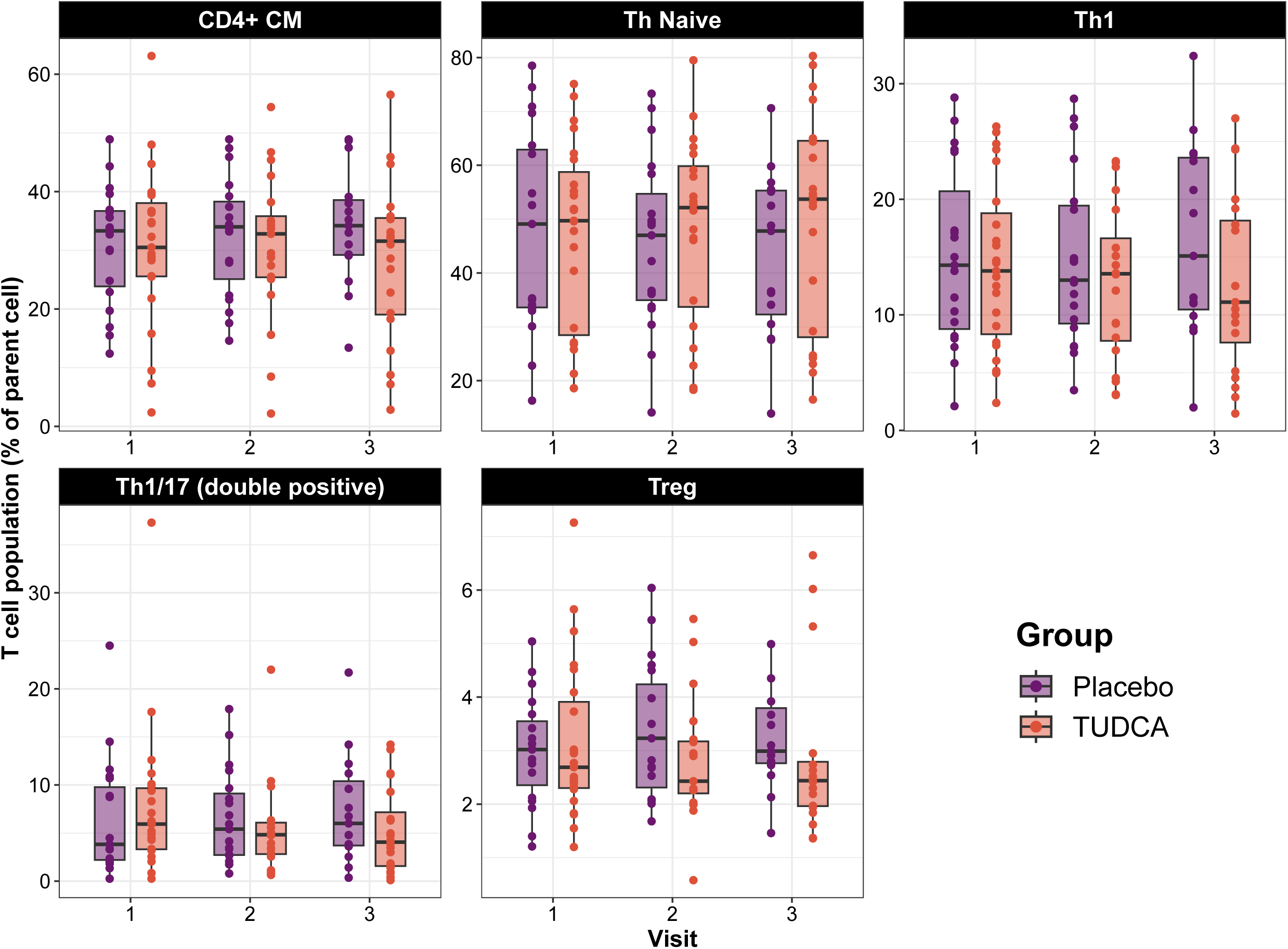
T cell population change over time in the clinical trial. Box plots overlapped by strip plots of T cell populations of the two groups throughout the study. Cells are expressed as a percent of their parent population. Bounds of the box represent interquartile range (IQR), horizontal central lines denote the median, while minimum and maximum whiskers correspond to the Q1−1.5×IQR (or the minimum value, if larger) and Q3+1.5×IQR (or the maximum value, if smaller), respectively. CM = central memory, Treg = regulatory T cells

**Table 7.**
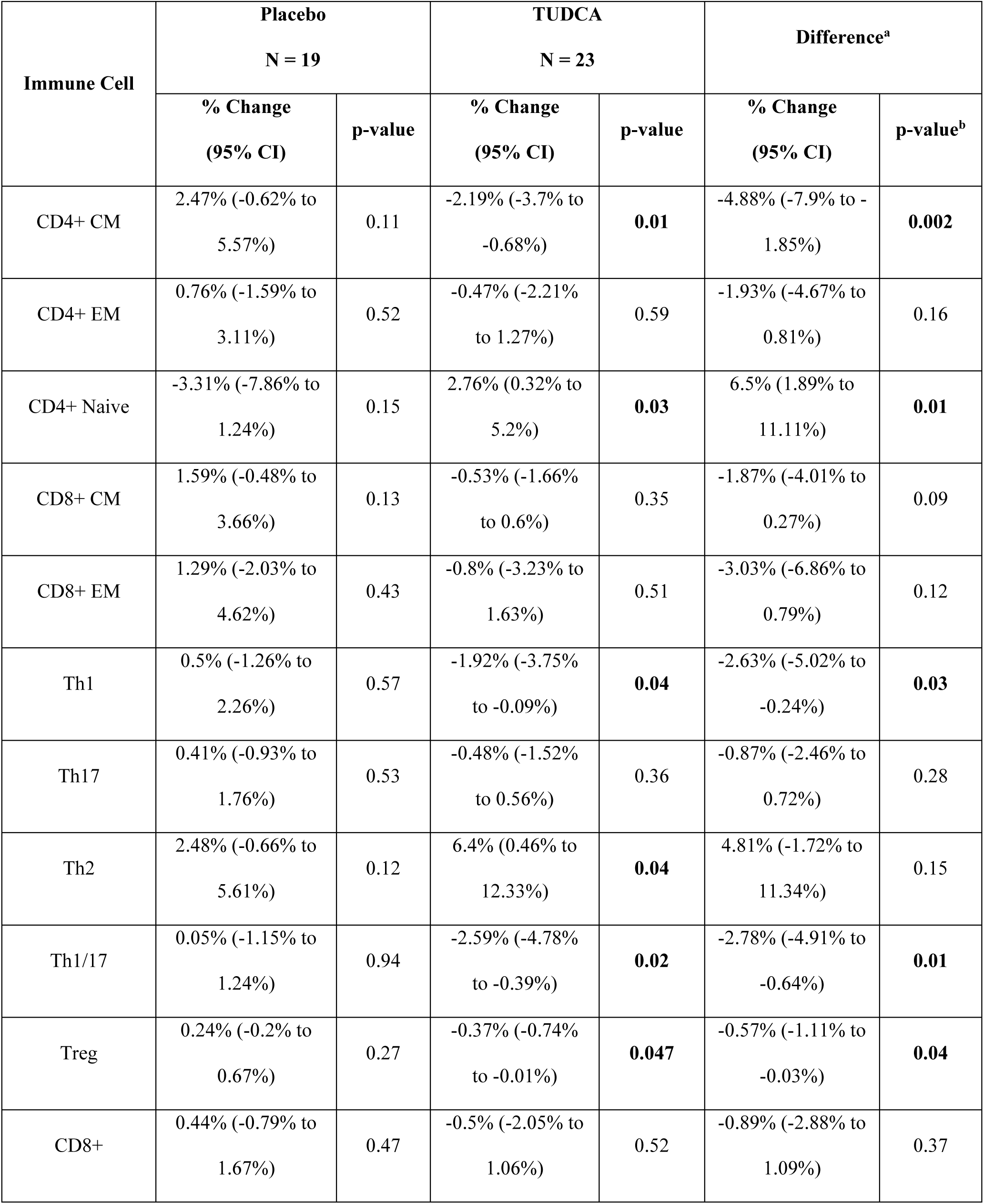
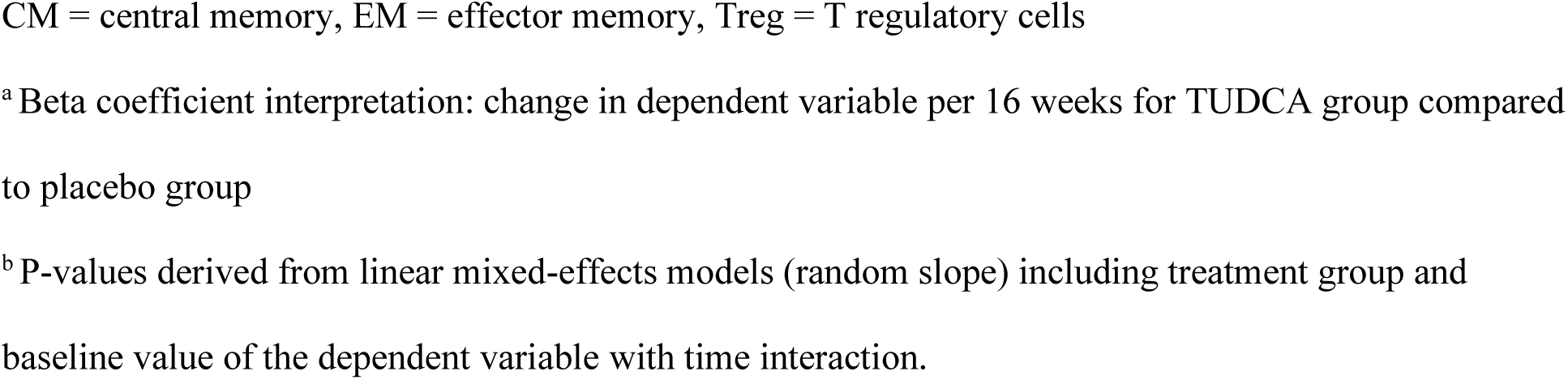
Comparison of T Cell Subset Changes (change per 16 weeks)

| Immune Cell | Placebo<br>N = 19 |  | TUDCA<br>N = 23 |  | Difference<sup>a</sup> |  |
| --- | --- | --- | --- | --- | --- | --- |
|  | % Change<br>(95% CI) | p-value | % Change<br>(95% CI) | p-value | % Change<br>(95% CI) | p-value<sup>b</sup> |
| CD4+ CM | 2.47% (-0.62% to 5.57%) | 0.11 | -2.19% (-3.7% to -0.68%) | **0.01** | -4.88% (-7.9% to -1.85%) | **0.002** |
| CD4+ EM | 0.76% (-1.59% to 3.11%) | 0.52 | -0.47% (-2.21% to 1.27%) | 0.59 | -1.93% (-4.67% to 0.81%) | 0.16 |
| CD4+ Naive | -3.31% (-7.86% to 1.24%) | 0.15 | 2.76% (0.32% to 5.2%) | **0.03** | 6.5% (1.89% to 11.11%) | **0.01** |
| CD8+ CM | 1.59% (-0.48% to 3.66%) | 0.13 | -0.53% (-1.66% to 0.6%) | 0.35 | -1.87% (-4.01% to 0.27%) | 0.09 |
| CD8+ EM | 1.29% (-2.03% to 4.62%) | 0.43 | -0.8% (-3.23% to 1.63%) | 0.51 | -3.03% (-6.86% to 0.79%) | 0.12 |
| Th1 | 0.5% (-1.26% to 2.26%) | 0.57 | -1.92% (-3.75% to -0.09%) | **0.04** | -2.63% (-5.02% to -0.24%) | **0.03** |
| Th17 | 0.41% (-0.93% to 1.76%) | 0.53 | -0.48% (-1.52% to 0.56%) | 0.36 | -0.87% (-2.46% to 0.72%) | 0.28 |
| Th2 | 2.48% (-0.66% to 5.61%) | 0.12 | 6.4% (0.46% to 12.33%) | **0.04** | 4.81% (-1.72% to 11.34%) | 0.15 |
| Th1/17 | 0.05% (-1.15% to 1.24%) | 0.94 | -2.59% (-4.78% to -0.39%) | **0.02** | -2.78% (-4.91% to -0.64%) | **0.01** |
| Treg | 0.24% (-0.2% to 0.67%) | 0.27 | -0.37% (-0.74% to -0.01%) | **0.047** | -0.57% (-1.11% to -0.03%) | **0.04** |
| CD8+ | 0.44% (-0.79% to 1.67%) | 0.47 | -0.5% (-2.05% to 1.06%) | 0.52 | -0.89% (-2.88% to 1.09%) | 0.37 |

CM = central memory, EM = effector memory, Treg = T regulatory cells
<sup>a</sup> Beta coefficient interpretation: change in dependent variable per 16 weeks for TUDCA group compared to placebo group
<sup>b</sup> P-values derived from linear mixed-effects models (random slope) including treatment group and baseline value of the dependent variable with time interaction.

### TUDCA supplementation alters the gut microbiome

To assess changes in the gut microbiota composition we performed shotgun metagenomics analysis on stool samples obtained at each study visit to define and quantify bacterial taxa and their functional profile. There were 34 participants that provided stool samples and were included in the analyses (13 in placebo arm and 21 in TUDCA arm). There was no significant change in the diet of the participants throughout the study, as assessed by the healthy eating index (HEI). TUDCA supplementation did not affect the alpha-diversity (a metric for the number of different species within a sample) as measured by Shannon index over time compared to placebo (Figure 3A). There was no difference in beta-diversity (the variance of microbiota between different samples) between the follow-up and baseline visits of the TUDCA group participants as measured by Bray-Curtis dissimilarity (p=0.7), but we noted a difference between TUDCA follow-up and placebo follow-up visits (p=0.002; Figure 3B). Relative abundances of taxa were derived from MetaPhlan, while relative abundances of functional features were derived from HUMAnN. Taxa analysis was mainly focused at the species level. Compared to placebo, 22 species demonstrated a decrease in abundance, while seven species demonstrated an increase in the TUDCA arm over the study period time (Figure 3C). All the individual species results can be found in Supplemental Table 3. Additionally, 14 metabolic pathways showed a downregulation over the study period in the TUDCA arm compared to placebo, while one pathway demonstrated upregulation (Figure 3D). We identified three pathways related to bile acids (bile acids epimerization, bile acid 7-alpha-dehydroxylation and bile acid 7-beta-dehydroxylation) but they did not demonstrate a difference between the two groups. All the functional analysis results are shown in Supplemental Table 4. Overall, these data demonstrate that TUDCA supplementation alters the composition and function of the gut microbiota in PMS.

**Figure 3.**
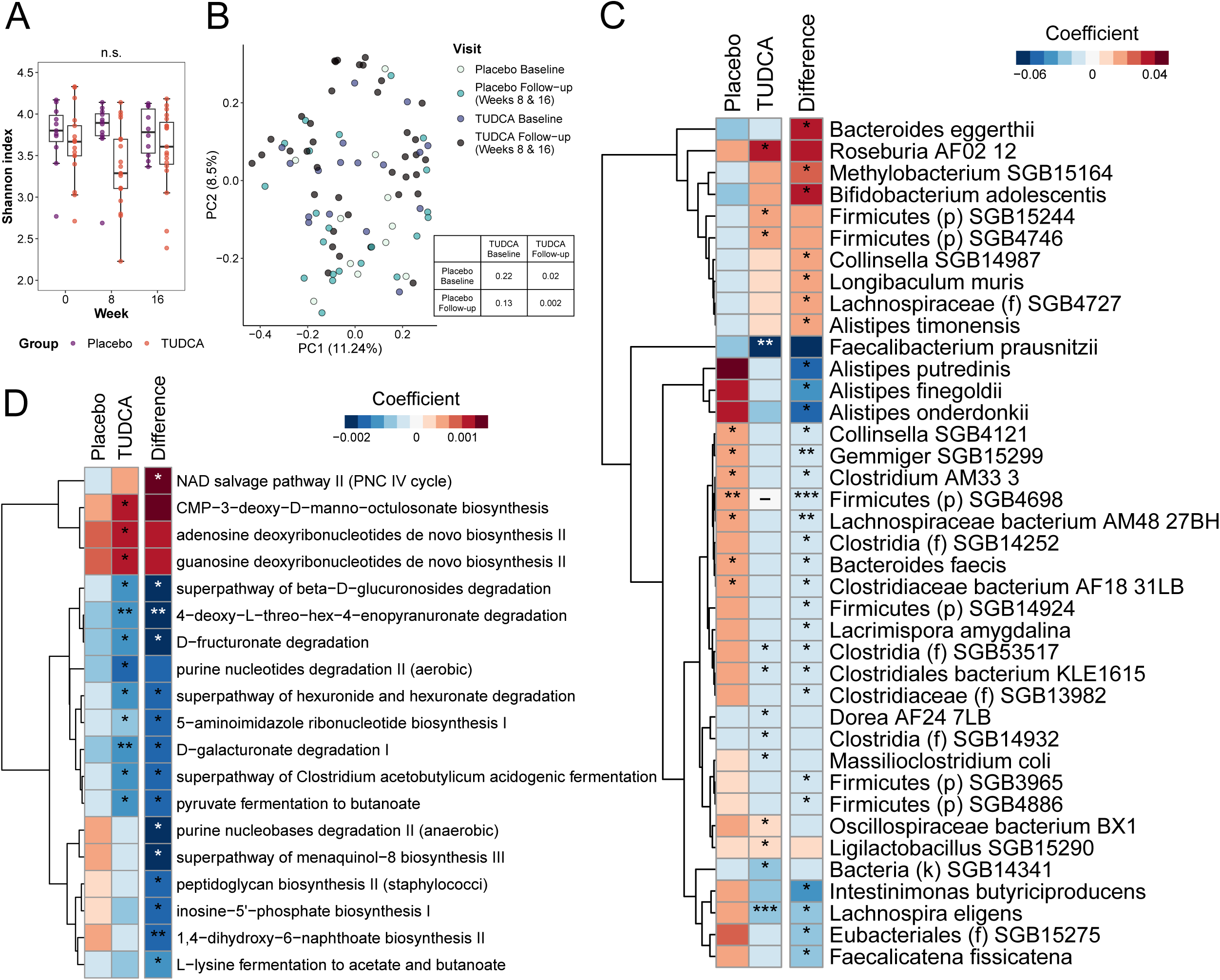
Changes in gut microbiome in the clinical trial. **(A)** Microbiome alpha-diversity measured by Shannon index in TUDCA and placebo (linear model). **(B)** PCoA of Bray-Curtis dissimilarity by group and visit (follow-up visits clustered together) demonstrating beta-diversity; table demonstrates pairwise p-values obtained from PERMANOVA. **(C and D)** Microbiota species **(C)** and pathways **(D)** that changed over time in TUDCA independently, as well as with comparison with placebo group (difference). *p<0.05, **p<0.01, ***p<0.01

## DISCUSSION

Building upon prior work that identified abnormal bile acid metabolism in MS, here we demonstrate in an observational cohort of people with MS that baseline circulating levels of bile acid metabolites predict subsequent disease progression as assessed using two imaging modalities – brain MRI and retinal OCT. Specifically, we noted that abnormal bile acid metabolism (particularly primary bile acids) predicted faster whole brain and brain substructure atrophy, in addition to predicting faster ONL atrophy in the retina. In our randomized, double-blind, placebo-controlled trial of TUDCA supplementation in people with PMS, we demonstrate the safety and tolerability of this intervention and that TUDCA supplementation increases circulating levels of multiple bile acids, impacts the peripheral immunophenotype and modifies the gut microbiome, as compared to placebo. Overall, our results provide additional data regarding the importance of bile acid metabolism in people with MS and rationale for future studies examining TUDCA supplementation or other interventions that target this metabolic pathway in MS.

Because our initial study had demonstrated that bile acid levels were altered in pediatric-onset and adults with MS, with more severe alterations noted in progressive disease, the relationships noted in the current study between bile acid levels and imaging outcomes were not surprising. Additionally, the presence of bile acid receptors, both nuclear (such as FXR) and on the cell surface (such as GPBAR1), on immune and glial cells in the brain and especially in MS lesions also provide a plausible mechanism by which circulating bile acids can impact disease progression. Our findings are congruent with a recent study that found lower circulating levels of primary bile acids and their precursor were associated with faster brain atrophy and more white matter lesion accumulation in aging (Baltimore Longitudinal Study of Aging; BLSA) and Alzheimer’s disease (Alzheimer’s Disease Neuroimaging Initiative; ADNI) cohorts (15). Other studies in mild cognitive impairment and Alzheimer’s disease have also shown altered bile acid metabolism and linked levels of specific bile acid metabolites/ ratios to specific cortical thickness measures (16). While we saw links of primary bile acid metabolism with whole brain and subcortical grey matter, there was no relationship noted with cortical white or grey matter atrophy. Whether this is related to the distribution of bile acid receptors in brain and retinal tissue is unknown, but could be a potential explanation for the relationships noted in our study. Unlike people with RRMS in whom the inner retina (RNFL and GICPL) is the predominant site of atrophy, in PMS we previously reported that the deep retina, specifically the ONL, atrophies faster than in RRMS (17). Given that bile acid metabolism is more altered in PMS (2), the above may provide a rationale for the observed relationship of primary bile acid metabolism specifically with ONL atrophy.

Consistent with previous bile acid supplementation trials (10, 14), TUDCA supplementation did not raise significant safety concerns and the TUDCA arm demonstrated a similar adverse event profile as the placebo arm in our trial. The gastrointestinal system was most commonly affected: diarrhea, nausea/vomiting and abdominal cramping were the most frequent adverse events, 12occurring in a higher frequency compared to placebo. On the other hand, the placebo group presented more frequently with non-GI events. One participant receiving TUDCA supplementation had a serious adverse event (cholangitis) leading to hospitalization, but it resolved without sequelae and TUDCA was considered an unlike cause by the DSMB. The participant had evidence of chronic changes in the biliary tree that predisposed to this condition and predated participation in the study. This event may however underscore the importance of excluding people with known abnormalities in the biliary tree anatomy from future TUDCA trials. Our results are congruent with those noted in both the pilot and the phase 2 randomized placebo-controlled trial of sodium phenylbutyrate and TUDCA supplementation in ALS, in which TUDCA was deemed to be safe and tolerable with mild gastrointestinal events occurring frequently (10, 14).

TUDCA supplementation resulted in significant increases in bile acids in the peripheral circulation. In the treatment group, UDCA, TUDCA, GUDCA, LCA and GLCA demonstrated an increase in their levels following treatment. All of these components are derivatives of the chenodeoxycholic acid (CDA) pathway. Naturally, unconjugated CDCA is metabolized by gut bacteria into UDCA or LCA. These secondary bile acids are reabsorbed and can become their conjugated (with either taurine or glycine) counterparts in the liver. When compared to placebo only changes in UDCA, TUDCA and GUDCA remained significant and given their closer proximity to TUDCA these results are not surprising. These bile acids have consistently demonstrated anti-inflammatory, anti-oxidant and anti-apoptotic properties in models of neurodegeneration (18).

TUDCA supplementation did not demonstrate significant effects on clinical or quality of life outcomes including EDSS, MSFC and MSQOL-54. Even though both EDSS and MSFC are validated MS disability measures and have been widely used (19), clinical disability progression based on these metrics needs to be confirmed ≥24 weeks apart, which extends beyond our study duration (20). These tools also lack the sensitivity to capture small amounts of change, especially in smaller sized cohorts. While we did note a significant improvement in the MSFC score in the TUDCA arm, this was not significantly different compared to the placebo arm. Change in patient-reported quality of life as measured by MSQOL-54 physical and mental components also did not differ between the two arms. Collectively, it is likely that a larger follow-up trial with a longer duration would be necessary to accurately assess the effects of TUDCA supplementation on clinical measures of disease severity.

Serum NfL and GFAP have emerged in recent years as biomarkers of neuroaxonal damage and astrogliosis, respectively (21, 22). In MS they are correlated with disease progression, clinical disability and brain atrophy and have been used as an adjunct to clinical and imaging measurements (23–25). The effect of DMT on serum NfL levels in people with PMS was assessed in the phase 3 placebo-controlled trial of ocrelizumab, where a significant decrease in NfL levels compared to placebo was observed at week 48 (26), which is three times longer than our study duration and hence it was not surprising to not observe a change in NfL levels over the course of our study. Additionally, two other two-year-long clinical trials in PMS failed to demonstrate a change in NfL levels, even when a reduction in the brain atrophy rate was achieved by the interventions (27–30). Data on how DMTs or other treatments affect serum GFAP are not yet available and hence similar to the clinical outcomes, a longer duration of treatment may perhaps have a greater chance of demonstrating an impact on these fluid biomarkers.

We performed immunophenotyping and noted that multiple T cell populations decreased significantly in the TUDCA supplementation group compared to placebo. Specifically, the proportion of central memory CD4+, Th1, Th1/17 (double positive) and Treg cells decreased, while naïve CD4+ T cells increased. In a study of mouse models of uveitis, supplementation of secondary bile acids resulted in reduced Th1 and Th17 cells in the spleen, as well as less severe inflammation in the uvea (31). These results were mediated through the GPBAR1. FXR – a bile acid-binding nuclear receptor – can also play an important role in T cell regulation. Hang et al showed that metabolites of LCA – a secondary bile acid –inhibited Th17 and promoted Treg differentiation in mice (32). Supplementation with LCA metabolites in mice decreased the number of ileal Th17 cells and protected mice from development of colitis (32). In our study LCA was significantly increased in the TUDCA arm and showed a trend towards an increase compared to the placebo arm, which could be one of the mechanisms underlying the observed T cell sub-population changes. Even though Tregs were reduced in our study, the changes in multiple different bile acids (although mostly secondary) and the alterations in the gut microbiota might have caused this finding. Overall, the reduction in multiple pro-inflammatory T cell populations likely overshadows the small reduction in Tregs.

We also performed profiling of the gut microbiota and observed changes in microbiota composition between the two trial arms. In total there were 29 species that changed their abundances in the TUDCA group compared to placebo during the 16-week study period. Of note, *Bifidobacterium adolescentis*, which has been found to be an avid gamma-aminobutyric acid (GABA) producer was one of the seven species that increased in the TUDCA treatment group (33, 34). GABA-producing gut bacteria can modulate the gut-brain axis response and therefore could potentially play a protective role in MS. Additionally, *B. adolescentis* abundance was found to be higher in centenarians and young individuals compared to an elderly cohort, suggesting it might have a beneficial effect in aging-related and degenerative changes (35). Four *Alistipes* species showed a change: *A. putredinis*, *A. finegoldii* and *A. onderdonkii* abundances decreased, and *A. timonensis* abundance increased over time in the TUDCA group compared to placebo. *Alistipes* is a relatively recently discovered genus that has been associated with multiple diseases, both in the gut and systemically (36). It has been associated with mood disorders with increasing abundance being associated with worsening symptoms, including fatigue and stress (37–39). Even though the particular species were not identified in these studies there might be an interplay of these bacteria with CNS symptoms. Furthermore, *Alistipes* species are indole-producing bacteria and can therefore affect tryptophan metabolism which was found to be altered in both pediatric and adult MS (40, 41). The remaining microbiota changes we observed in this study have not been previously studied in the literature, though many of them could not be characterized to the species levels (unknown species). These changes could be attributed to the fact that bile salts might be toxic to bile-sensitive bacteria (42).

In addition to changes in the abundance of gut bacteria, changes in microbiota metabolic function are also important. We identified 15 different pathways that changed with TUDCA supplementation. Interestingly, NAD salvage pathway II (PNC IV cycle) was upregulated in the TUDCA arm compared to placebo. NAD is an energy component with an important role in numerous processes including but not limited to redox homeostasis, inflammation, mitochondrial homeostasis, metabolism, gene expression and circadian rhythm (43). Depletion of NAD can deregulate these fundamental functions leading to mitochondrial dysfunction, increased inflammation and accelerated ageing. NAD depletion has been linked to many neurodegenerative diseases and NAD boosting has been used as a therapeutic strategy (43). Gut bacteria have been shown to play an important role in the liver – and subsequently systemic – NAD pool as well (44). Therefore, upregulation of NAD salvage pathway in the gut microbiota could potentially lead to increased NAD levels in the periphery and consequently in the CNS. On the other hand, there were 14 pathways that showed a downregulation in the TUDCA group compared to control. Beta-glucuronidase plays a role in the elimination of multiple endobiotics and xenobiotics (45). It is involved in drug metabolism and can affect the blood estrogen pool. Whether it can play a role in inflammation or CNS function is unknown. The other downregulated pathways are mainly involved in dietary fiber breakdown (e.g. D−fructuronate degradation, D−galacturonate degradation I) and bacterial nucleotide synthesis (e.g. 5−aminoimidazole ribonucleotide biosynthesis I, inosine−5’−phosphate biosynthesis I, purine nucleobases degradation II). Overall, we noted multiple changes in the gut microbiota with TUDCA supplementation several of which could have biologically relevant effects in people with MS.

There were certain limitations of this study. The observational cohort used for assessing the baseline metabolite levels was identified retrospectively and hence we focused on objective measures of disease progression including MRI and OCT imaging. Additionally, for this cohort we did not have fasting data for the participants. For the clinical trial, the short study duration and small sample size may have limited the ability to capture significant effects of TUDCA supplementation on clinical and fluid biomarker outcomes. Also, there were no imaging outcomes assessed in this study limiting the ability to evaluate TUDCA’s effects on structural components of disease activity and progression in the brain and spinal cord, although 16 weeks would likely be too short to assess meaningful changes. Lastly, the effects of the COVID-19 pandemic impacted trial activities, necessitating that some visits be conducted via telemedicine rather than in-person as previously planned, and resulting in some missing samples and data points, as well as some participants being lost to follow up.

In conclusion, we show that circulating bile acid metabolites predict MS disease progression assessed using imaging outcomes. Oral TUDCA supplementation is safe and tolerable in people with progressive MS and results in increased circulating bile acid levels, altered peripheral immunophenotype and the gut microbiota. Future studies with a larger sample size and longer duration are needed to validate our findings and to better assess effects of TUDCA supplementation on clinical and imaging outcomes in people with MS.

## METHODS

### Sex as a biological variable

Our study examined males and females, and similar findings are reported for both sexes.

### Observational cohort

#### Study population, serum metabolomics and imaging data

People with MS with a serum specimen within 12 months interval of a brain MRI scan and retinal OCT scan were identified. Those with serial brain MRI (annually) and retinal OCT (6-monthly) scans were included. Upon collection, blood specimens were centrifuged, and serum was isolated and stored in -80°C. Specimens were sent frozen to Metabolon for global metabolomic analysis, as previously described (46). MRI and OCT images were segmented by in-house segmentation algorithms and images underwent quality control manually (47–50); images with failed segmentations or artifacts were excluded. Whole brain volume and brain substructure volumes were derived and adjusted for individualized intracranial volume. Detailed methods about MRI and OCT scan acquisition and segmentation, are the same as published before (51).

#### Statistics

We utilized log-linear mixed-effects models with normalized whole brain and brain substructure volumes and retinal thicknesses as the dependent variables and individual metabolites with time (modeled as years from baseline) interaction as the independent variables. All models included subject-specific random intercepts and slopes, while the ones with retinal layers included additionally eye-specific random intercepts and slopes. Analyses were adjusted for age, sex, and MS subtype with time interaction. Models with retinal layers as the outcome were additionally adjusted for eye-specific history of optic neuritis and eyes with active optic neuritis episodes during follow-up were excluded. We then performed metabolite set enrichment analysis (MSEA), which, similar to gene set enrichment analyses, uses the test statistics from the individual metabolite analysis, as described elsewhere (46) as well as pre-specified metabolite groupings (based on biological classification from Metabolon). MSEA then tests whether statistically significant similar change in metabolite levels occur within each group; NESs and associated p-values were calculated as summary measures of the effect size, directionality and statistical significance of concordant changes within groups. Since our study was focused on bile acids, only results from the primary and secondary bile acid metabolism pathways were included.

### Clinical Trial

#### Study Design and Procedures

This was a randomized, double-blind, placebo-controlled, single-center phase 1/2a trial conducted at Johns Hopkins University from June 2018 to April 2022 evaluating the safety and efficacy of TUDCA supplementation in progressive MS. People with PMS fulfilling eligibility criteria were enrolled after a screening period of at least two weeks, and randomized (1:1) to receive TUDCA 1g twice daily or placebo for a duration of 16 weeks. Randomization was conducted by the study statistician from Johns Hopkins School of Public Health. All participants and study personnel, except the study statistician, were blinded to the study treatment.

Four scheduled study visits were performed as follows - screening, 0 weeks (baseline), 8 weeks (mid-study), and 16 weeks (end-of-study). Due to the COVID-19 pandemic, the screening and baseline visits were later combined into one visit in order to limit the in-person visits.

Furthermore, on occasions that the participant could not perform an in-person visit, a remote tele-visit was held instead. Verification of eligibility was performed by the treating physician and baseline demographics were collected at the screening visit. Assessments including vitals with physical and neurological examinations, labs, urine pregnancy tests (if applicable), suicidality evaluations, and review of medical history and current medications were conducted at each visit. Assessment of adverse events, review of safety lab results, and collection of blood including peripheral blood mononuclear cells (PBMC) for peripheral immunophenotyping were performed at 0 weeks, 8 weeks, and 16 weeks. Blinded Multiple Sclerosis Functional Composite (MSFC) and Multiple Sclerosis Quality of Life-54 (MSQOL-54) assessments were performed at all visits. Blinded Expanded Disability Status Scale (EDSS) evaluations were performed at 0 weeks, 8 weeks, and 16 weeks. Food frequency questionnaire assessments and stool specimens were collected at 0 weeks, 8 weeks, and 16 weeks. Participants were returning the pill bottles at weeks 8 and 16, and adherence was documented by the study pharmacist.

#### Inclusion and exclusion criteria

Eligibility criteria included: adults age ≥18; diagnosis of progressive MS according to Lublin criteria (52); on the same disease modifying therapy for ≥ 6 months and not expected to change therapy in the next 6 months; and no relapse in the prior 3 months. Exclusion criteria were a known history of other neuroinflammatory, neurodegenerative or systemic autoimmune disease; prior history of liver disease; stage IV/V chronic kidney disease or other severe metabolic derangements; BMI < 15 kg/m^2^ and BMI > 40 kg/m^2^; females who were pregnant, nursing, or not willing to use contraception; chronic antibiotic use; corticosteroid treatment within the past 30 days; and history of bariatric surgery, small intestinal surgery resulting in loss of length of the jejunum or ileum, chronic disease of the small intestine (e.g., Crohn’s Disease, gluten sensitive enteropathy, or other disorders that could lead to malabsorption), abnormal bile duct anatomy, sclerosing cholangitis, or positive anti-mitochondrial antibodies.

#### Safety labs

Safety labs were drawn in each study visit and included serum aspartate transaminase (AST), alanine transaminase (ALT), bilirubin, creatinine, and a urine pregnancy test if applicable.

Participants would discontinue treatment if in case of abnormal safety labs defined as serum AST or ALT >3x upper limit of normal, serum bilirubin >3x upper limit of normal, estimated glomerular filtration rate (eGFR) <60ml/min/1.73m^2^ or positive pregnancy test during the study.

#### Outcomes

The primary outcome of this trial was the safety and tolerability of TUDCA supplementation, which was assessed by the number of adverse events and rate of drop out in each group.

Exploratory secondary outcomes assessed were changes in the following - metabolomics using bile acid serum levels, clinical scores (EDSS, MSFC, and MSQOL-54), NfL and GFAP levels, gut microbiota and peripheral immune cells. EDSS and MSFC assessments were performed by trained personnel, blinded to treatment status. The MSQOL-54 and NutritionQuest dietary questionnaires were provided to the participants along with a pre-labeled mailing package, which they used to mail them back. The validated 2014 Block Food Frequency Questionnaire and Physical Activity Screener (NutritionQuest) was administered throughout the study.

Questionnaires assessing the past year were used during the baseline visit, while past-month questionnaires were used at consecutive study visits.

#### Blood samples

Plasma was isolated from the blood samples and was stored at -80°C until use. 1ml samples were shipped to Metabolon (Durham, NC, USA) in dry ice for targeted bile acid analysis using liquid chromatography and mass spectrometry. This targeted panel measured 15 major human primary and secondary bile acids and their conjugates. NfL and GFAP were quantified in the serum using the Simoa neurology 2-plex b (NfL, GFAP) assay (Quanterix, Billerica, MA), according to the manufacturer’s instructions. Samples belonging to the same subject were run on the same plate.

#### Stool sample collection and Illumina shotgun sequencing

Participants were provided with a stool sample collection kit (OMNIgene GUT OMR-200; DNA Genotek Inc., Ottawa, Ontario, Canada) and instructed how to collect the sample in their home. The samples were returned and stored at -80°C. Once the study was completed, all collected stool kits were shipped frozen in dry ice to MicrobiomeInsights (Vancouver, British Columbia, Canada) for further analysis. DNA libraries were prepared with the Nextera DNA Flex kit (Illumina Inc., San Diego, CA, USA) using an in-house protocol. Sequencing was done on an Illumina NovaSeq (150 bp x 2).

#### Whole metagenome shotgun sequencing and data processing

Demultiplexed shotgun metagenomic paired sequences were processed with Kneaddata (v.0.10.0; default parameters) for trimming (removal of low-quality bases, reads and adapters) using Trimmomatic (53) and for decontamination from human DNA using BowTie2 (54) to map the reads against the human reference database (hg37_and_human_contamination).

#### Taxonomic and functional profiling

MetaPhlAn 4 (v.4.0.6; default parameters) was used for taxonomic profiling (55). Briefly, MetaPhlAn is a computational tool that maps shotgun metagenomics reads to species-level. MetaPhlAn 4 relies on unique clade-specific marker genes identified from microbial genome reference database. In a sensitivity analysis, taxonomy was derived using Kraken2 with the PlusPF database, which contains bacterial, archaeal, micro-eukaryotic, and viral genomes from Refseq (56). Functional analysis was performed utilizing HUMAnN 3 (v.3.7; default parameters) (57). HUMAnN provides the abundance of metabolic pathways by bacterial species, based on numerous available databases, including KEGG, GO, Metacyc, and Level-4 enzyme commission.

#### PBMC Isolation

PBMCs were isolated from whole blood in sodium heparin coated vacutainers using SepMate PBMC Isolation Tubes with LymphoPrep density gradient medium. Briefly, whole blood was mixed with room temperature PBS 1:1, then up to 34 mL (17mL blood) was overlayed on top of 15 mL density gradient medium in a 50mL SepMate tube. Tubes were spun 1200g for 10 minutes at room temperature then approximately 10mL of plasma was removed and the remaining plasma and PBMCs were decanted into a new 50mL centrifuge tube. Cells were washed in chilled PBS twice then resuspended in 1mL complete IMDM (IMDM with 10% human serum, 4 mM L-Glutamine, 1000U/mL Penicillin, 1000µg/mL streptomycin, 5mg/mL gentamicin) and counted using Acridine Orange + Propidium Iodide (AO/PI) on a Countess 3 automated hemocytometer. Volume was adjusted with cIMDM to reach approximate cell concentration of 2e7 cells/mL and mixed 1:1 with 20% DMSO in cIMDM for a final concentration of approximately 1e7 cells/mL in 10% DMSO cIMDM. Cells were aliquoted in 1mL volume in cryovials and frozen overnight at a controlled rate using Mr. Frosty freezing container. The following day (or Monday for cells isolated on Friday), cells were moved to vapor phase liquid nitrogen for long term storage.

#### Flow Cytometry

Flow cytometry was performed across 4 days, with all visits from an individual being done on the same day, and each day containing a mix of vehicle and treatment participants. PBMC aliquots were retrieved from liquid nitrogen and incubated at 37C for approximately 6 minutes in a ThermoMixer C with cryo thaw adapter. Thawed cells were then poured into 15mL conicals containing pre-warmed 50% FBS in PBS with 100U/mL DNase I. Tubes were capped and inverted gently. Cells were spun for 10 minutes at 300g then supernatant was decanted. Cell pellet was disrupted by running tube over 15mL tube holding rack and resuspended in 500ul PBS. 10uL were removed and counted using AO/PI. 5e5 cells were transferred into 96 well V-bottom microplates and the volume was brought up to 250ul in PBS. Plates were spun at 500g and supernatant decanted. Cells were then incubated in PBS with 1:2000 Zombie NIR permeability stain (Biolegend) and 1:20 TruStain FcX Blocker antibody (Biolegend) for 15 minutes at room temperature. Plates were spun again at 500g, supernatant decanted, and cells were stained in 50ul surface marker antibodies (see Supplemental Table 5) in FACS buffer (2% FBS in PBS with 2mM EDTA) and incubated at room temperature for 30 minutes. Cells were washed in 200ul FACS buffer, centrifuged, then resuspended in 200ul FoxP3 Fixation/Permeabilization Buffer (Thermo Fisher/eBioscience) and incubated for 20 minutes at room temperature. Cells were centrifuged and washed in 200ul Permeabilization Buffer (Thermo Fisher/eBioscience), centrifuged again, then stained for intracellular markers in 50ul Permeabilization Buffer and incubated for 1 hour at room temperature. Cells were then washed in permeabilization buffer, centrifuged, washed in FACS buffer, centrifuged, resuspended in 100ul FACS buffer and analyzed on Cytek Aurora 3L (VBR) Spectral Cytometer. Analysis of cytometry data was done in FlowJo (v10.9.0) with manual gating using fluorescence-minus-one (FMO) controls as needed. Gating strategy and relevant FMO controls are shown in Supplemental Figure.

#### Statistics

For all the analyses, a modified intention-to-treat approach was followed and only participants that had at least one follow-up visit were included.

Incidence rate of adverse events was recorded and summarized by system organ class, severity, and relationship to study treatment. Safety data for both groups was compared over the 16-week treatment period and was analyzed using Fisher’s exact test.

The difference in the change over time in each of the outcomes (metabolomics, clinical measures, biomarkers, microbiome functional outcomes, peripheral immune cells) between the two groups were calculated using linear mixed-effects models with the outcome as the dependent variable and the treatment group, time and their interaction as the independent variables. Additionally, all analyses besides those for microbiome outcomes were adjusted for the baseline measurement of the outcome (and their interaction with time) to account for any differences at baseline. The rate of change in the relative abundance of the bacteria species was calculated using zero-inflated gaussian mixed-effects models due to the sparse nature of the data (58, 59). Mixed-effects models included participant-specific random intercepts and slopes assuming model convergence. If the model did not converge, random slopes were dropped. Only subjects with at least two measurements for each outcome were included in the analyses.

Metabolomics data for changes in serum bile acid levels between the groups over the 16-week period was assessed at the end of the study using quantitation of 15 bile acids included in our panel. Undetected levels were set half of the minimal detection value. The data was log-transformed to approximate normal distribution.

Changes in serum NfL and GFAP levels over the 16-week period were assessed for both groups using linear mixed-effects regression models to determine whether there was a difference between NfL and GFAP rates of change between the two groups. NfL and GFAP levels were log-transformed to approximate normality.

The peripheral immune cells were analyzed as a percentage of their parent populations.

Alpha (using Shannon index) and beta diversity (using Bray-Curtis dissimilarity) were calculated and the difference in rate of change in alpha-diversity between the two arms was calculated with linear mixed-effects models. The PERMANOVA (permutational multivariate ANOVA) test was used to compare beta diversity between two groups at baseline and follow-up visits using adonis2 function of vegan package in R (60). To deal with sparse microbial data in the analysis, we focused on species with relative abundance of at least 0.0001, in at least 5% of the samples. A total of 554 species were included. Zero inflated gaussian mixed-effects models were utilized to identify species that changed over time in the two groups (58, 59). The relative abundances of functional pathways were analyzed using linear mixed-effects models. We focused on pathways that were present in at least 5% of the samples (436 pathways). For these analyses, the relative abundance of microbial composition and pathway were transformed with an arcsine square root transformation (58).

In order to assess whether there was a significant change in the subjects’ dietary habits, the Healthy Eating Index was calculated from the 2014 Block Food Frequency Questionnaire and its rate of change across the study period was calculated with linear mixed-effects model.

All statistical analyses were performed using R Version 4.3.1 (https://www.r-project.org/). Statistical significance was defined as p<0.05.

### Study approval

Approval of the study protocol and procedures was obtained from Johns Hopkins University Institutional Review Board (IRB00144766). The trial was registered on clinicaltrials.gov (NCT03423121). Written informed consent was obtained from all participants at the screening visit.

## Data Availability

De-identified data are available from the corresponding author upon reasonable request.

## CONTRIBUTIONS

PB, PAC contributed to the study design. DCL, KLH, MDS, KS, SG, LJ, SH, FF, BED, KCF, ESS, SS, PB conducted experiments and contributed to the acquisition of data. DCL and PB contributed to the statistical analyses. DCL, KLH, MDS and PB contributed significantly to preparing the manuscript and figures. All authors contributed editing the final manuscript. The order for co-first authors was determined by flipping a coin.

## ACKNOWLEDGEMENTS

This study was supported by the National MS Society through a research grant to PB. LJ received funding through a postdoctoral fellowship from the National MS Society. We would like to thank Dr. Michael Levy, Dr. Kirti Shetty and Dr. Kathryn Fitzgerald for serving on the Data Safety Monitoring Board for the trial.

---

**Supplemental Figure.**
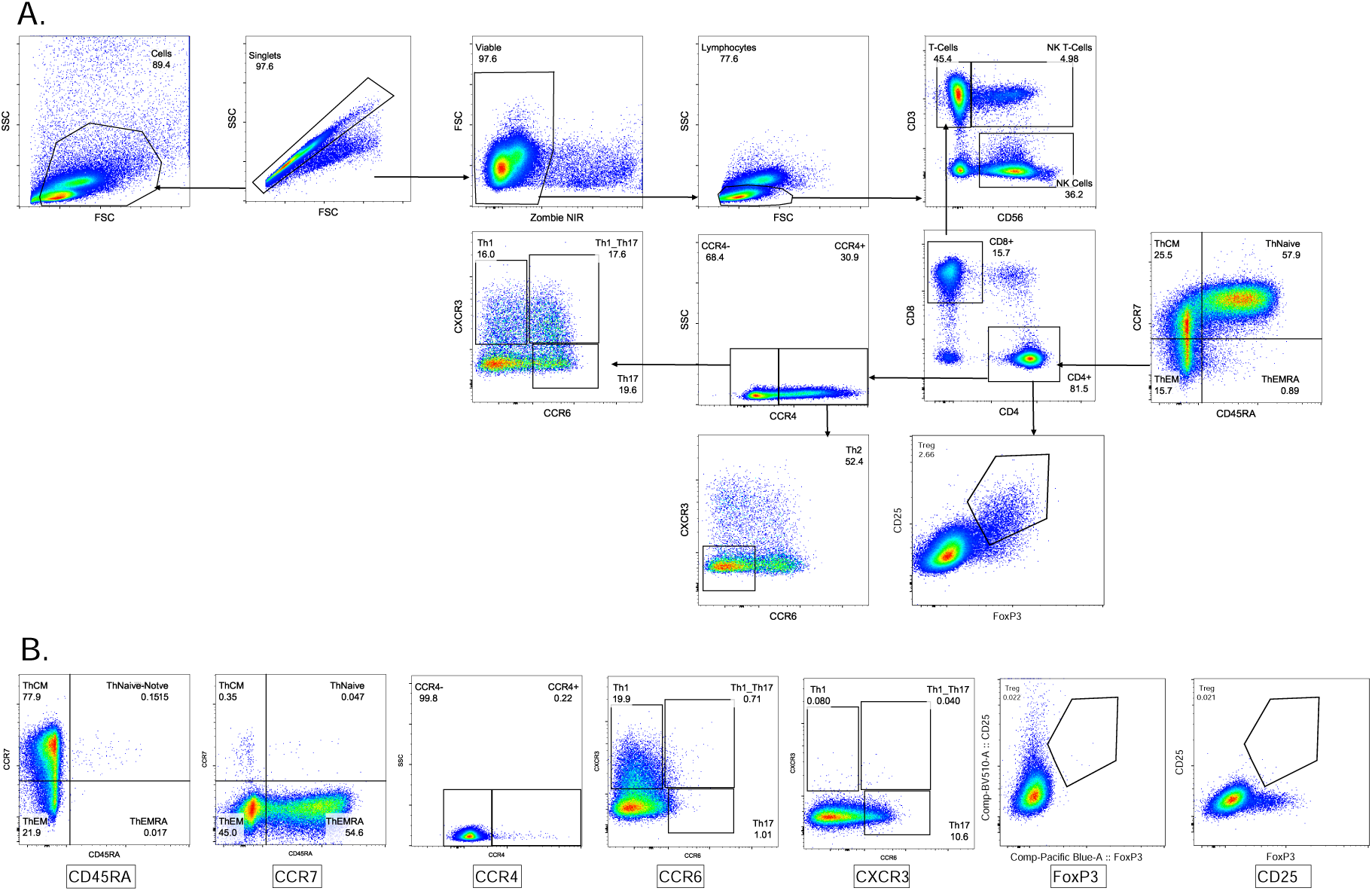
Gating strategy for flow cytometry analyses. **(A)** Representative gating strategy for subsetting flow cytometry data to quantify populations of interest. **(B)** Fluorescence-minus-one controls for relevant antibodies

## REFERENCES

1. Reich DS, Lucchinetti CF, Calabresi PA. Multiple Sclerosis. N Engl J Med. 2018;378(2):169– 180.

2. Bhargava P, et al. Bile acid metabolism is altered in multiple sclerosis and supplementation ameliorates neuroinflammation. J Clin Invest. 2020;130(7):3467–3482.

3. Miyake S, et al. Dysbiosis in the Gut Microbiota of Patients with Multiple Sclerosis, with a Striking Depletion of Species Belonging to Clostridia XIVa and IV Clusters. PloS One. 2015;10(9):e0137429.

4. Jangi S, et al. Alterations of the human gut microbiome in multiple sclerosis. Nat Commun. 2016;7:12015.

5. Chen J, et al. Multiple sclerosis patients have a distinct gut microbiota compared to healthy controls. Sci Rep. 2016;6:28484.

6. Castro-Caldas M, et al. Tauroursodeoxycholic Acid Prevents MPTP-Induced Dopaminergic Cell Death in a Mouse Model of Parkinson’s Disease. Mol Neurobiol. 2012;46(2):475–486.

7. Gaspar JM, et al. Tauroursodeoxycholic acid protects retinal neural cells from cell death induced by prolonged exposure to elevated glucose. Neuroscience. 2013;253:380–388.

8. Gómez-Vicente V, et al. Neuroprotective Effect of Tauroursodeoxycholic Acid on N-Methyl-D-Aspartate-Induced Retinal Ganglion Cell Degeneration. PLOS ONE. 2015;10(9):e0137826.

9. Keene CD, et al. Tauroursodeoxycholic acid, a bile acid, is neuroprotective in a transgenic animal model of Huntington’s disease. Proc Natl Acad Sci U S A. 2002;99(16):10671–10676.

10. Elia AE, et al. Tauroursodeoxycholic acid in the treatment of patients with amyotrophic lateral sclerosis. Eur J Neurol. 2016;23(1):45–52.

11. Min J-H, et al. Oral solubilized ursodeoxycholic acid therapy in amyotrophic lateral sclerosis: a randomized cross-over trial. J Korean Med Sci. 2012;27(2):200–206.

12. Parry GJ, et al. Safety, Tolerability, and Cerebrospinal Fluid Penetration of Ursodeoxycholic Acid in Patients With Amyotrophic Lateral Sclerosis. Clin Neuropharmacol. 2010;33(1):17.

13. Paganoni S, et al. Long-term survival of participants in the CENTAUR trial of sodium phenylbutyrate-taurursodiol in amyotrophic lateral sclerosis. Muscle Nerve. 2021;63(1):31–39.

14. Paganoni S, et al. Trial of Sodium Phenylbutyrate–Taurursodiol for Amyotrophic Lateral Sclerosis. N Engl J Med. 2020;383(10):919–930.

15. Varma VR, et al. Bile acid synthesis, modulation, and dementia: A metabolomic, transcriptomic, and pharmacoepidemiologic study. PLOS Med. 2021;18(5):e1003615.

16. Nho K, et al. Altered Bile Acid Profile in Mild Cognitive Impairment and Alzheimer’s Disease: Relationship to Neuroimaging and CSF Biomarkers. Alzheimers Dement J Alzheimers Assoc. 2019;15(2):232–244.

17. Sotirchos ES, et al. Progressive Multiple Sclerosis Is Associated with Faster and Specific Retinal Layer Atrophy. Ann Neurol. 2020;87(6):885–896.

18. Huang F, Pariante CM, Borsini A. From dried bear bile to molecular investigation: A systematic review of the effect of bile acids on cell apoptosis, oxidative stress and inflammation in the brain, across pre-clinical models of neurological, neurodegenerative and neuropsychiatric disorders. Brain Behav Immun. 2022;99:132–146.

19. Cohen JA, et al. Disability outcome measures in multiple sclerosis clinical trials: current status and future prospects. Lancet Neurol. 2012;11(5):467–476.

20. Cadavid D, et al. The EDSS-Plus, an improved endpoint for disability progression in secondary progressive multiple sclerosis. Mult Scler J. 2017;23(1):94–105.

21. Khalil M, et al. Serum neurofilament light levels in normal aging and their association with morphologic brain changes. Nat Commun. 2020;11(1):812.

22. Abdelhak A, et al. Blood GFAP as an emerging biomarker in brain and spinal cord disorders. Nat Rev Neurol. 2022;18(3):158–172.

23. Barro C, et al. Serum GFAP and NfL Levels Differentiate Subsequent Progression and Disease Activity in Patients With Progressive Multiple Sclerosis. Neurol Neuroimmunol Neuroinflammation. 2022;10(1):e200052.

24. Uphaus T, et al. NfL predicts relapse-free progression in a longitudinal multiple sclerosis cohort study. EBioMedicine. 2021;72:103590.

25. Abdelhak A, et al. Serum GFAP as a biomarker for disease severity in multiple sclerosis. Sci Rep. 2018;8(1):14798.

26. Bar-Or A, et al. Blood neurofilament light levels predict non-relapsing progression following anti-CD20 therapy in relapsing and primary progressive multiple sclerosis: findings from the ocrelizumab randomised, double-blind phase 3 clinical trials. eBioMedicine. 2023;93:104662.

27. Chataway J, et al. Effect of high-dose simvastatin on brain atrophy and disability in secondary progressive multiple sclerosis (MS-STAT): a randomised, placebo-controlled, phase 2 trial. The Lancet. 2014;383(9936):2213–2221.

28. Williams TE, et al. Assessing Neurofilaments as Biomarkers of Neuroprotection in Progressive Multiple Sclerosis. Neurol Neuroimmunol Neuroinflammation. 2022;9(2):e1130.

29. Fox RJ, et al. Phase 2 Trial of Ibudilast in Progressive Multiple Sclerosis. N Engl J Med. 2018;379(9):846–855.

30. Fox RJ, et al. Neurofilament light chain in a phase 2 clinical trial of ibudilast in progressive multiple sclerosis. Mult Scler J. 2021;27(13):2014–2022.

31. Hu J, et al. Gut microbiota-mediated secondary bile acids regulate dendritic cells to attenuate autoimmune uveitis through TGR5 signaling. Cell Rep. 2021;36(12):109726.

32. Hang S, et al. Bile acid metabolites control TH17 and Treg cell differentiation. Nature. 2019;576(7785):143–148.

33. Kali A. Psychobiotics: An emerging probiotic in psychiatric practice. Biomed J. 2016;39(3):223–224.

34. Duranti S, et al. Bifidobacterium adolescentis as a key member of the human gut microbiota in the production of GABA. Sci Rep. 2020;10(1):14112.

35. Wu L, et al. A Cross-Sectional Study of Compositional and Functional Profiles of Gut Microbiota in Sardinian Centenarians. mSystems. 2019;4(4):e00325–19.

36. Parker BJ, et al. The Genus Alistipes: Gut Bacteria With Emerging Implications to Inflammation, Cancer, and Mental Health. Front Immunol. 2020;11:906.

37. Frémont M, et al. High-throughput 16S rRNA gene sequencing reveals alterations of intestinal microbiota in myalgic encephalomyelitis/chronic fatigue syndrome patients. Anaerobe. 2013;22:50–56.

38. Naseribafrouei A, et al. Correlation between the human fecal microbiota and depression. Neurogastroenterol Motil. 2014;26(8):1155–1162.

39. Jiang H, et al. Altered fecal microbiota composition in patients with major depressive disorder. Brain Behav Immun. 2015;48:186–194.

40. Nourbakhsh B, et al. Altered tryptophan metabolism is associated with pediatric multiple sclerosis risk and course. Ann Clin Transl Neurol. 2018;5(10):1211–1221.

41. Fitzgerald KC, et al. Multi-omic evaluation of metabolic alterations in multiple sclerosis identifies shifts in aromatic amino acid metabolism. Cell Rep Med. 2021;2(10):100424.

42. Urdaneta V, Casadesús J. Interactions between Bacteria and Bile Salts in the Gastrointestinal and Hepatobiliary Tracts. Front Med. 2017;4:163.

43. Xie N, et al. NAD+ metabolism: pathophysiologic mechanisms and therapeutic potential. Signal Transduct Target Ther. 2020;5(1):1–37.

44. Shats I, et al. Bacteria Boost Mammalian Host NAD Metabolism by Engaging the Deamidated Biosynthesis Pathway. Cell Metab. 2020;31(3):564–579.e7.

45. Wang P, et al. Human gut bacterial β-glucuronidase inhibition: An emerging approach to manage medication therapy. Biochem Pharmacol. 2021;190:114566.

46. Virupakshaiah A, et al. Several serum lipid metabolites are associated with relapse risk in pediatric-onset multiple sclerosis. Mult Scler J. 2023;29(8):936–944.

47. Dewey BE, et al. DeepHarmony: A deep learning approach to contrast harmonization across scanner changes. Magn Reson Imaging. 2019;64:160–170.

48. Zhao C, et al. SMORE: A Self-Supervised Anti-Aliasing and Super-Resolution Algorithm for MRI Using Deep Learning. IEEE Trans Med Imaging. 2021;40(3):805–817.

49. Huo Y, et al. 3D whole brain segmentation using spatially localized atlas network tiles. NeuroImage. 2019;194:105–119.

50. Huo Y, et al. Consistent cortical reconstruction and multi-atlas brain segmentation. NeuroImage. 2016;138:197–210.

51. Murphy OC, et al. Trans-Synaptic Degeneration Following Acute Optic Neuritis in Multiple Sclerosis. Ann Neurol. 2023;93(1):76–87.

52. Lublin FD, et al. Defining the clinical course of multiple sclerosis. Neurology. 2014;83(3):278–286.

53. Bolger AM, Lohse M, Usadel B. Trimmomatic: a flexible trimmer for Illumina sequence data. Bioinformatics. 2014;30(15):2114–2120.

54. Langmead B, Salzberg SL. Fast gapped-read alignment with Bowtie 2. Nat Methods. 2012;9(4):357–359.

55. Blanco-Míguez A, et al. Extending and improving metagenomic taxonomic profiling with uncharacterized species using MetaPhlAn 4. Nat Biotechnol. 2023;1–12.

56. Wood DE, Lu J, Langmead B. Improved metagenomic analysis with Kraken 2. Genome Biol. 2019;20(1):257.

57. Beghini F, et al. Integrating taxonomic, functional, and strain-level profiling of diverse microbial communities with bioBakery 3. eLife. 2021;10:e65088.

58. Zhang X, Guo B, Yi N. Zero-Inflated gaussian mixed models for analyzing longitudinal microbiome data. PLoS ONE. 2020;15(11):e0242073.

59. Kodikara S, Ellul S, Lê Cao K-A. Statistical challenges in longitudinal microbiome data analysis. Brief Bioinform. 2022;23(4):bbac273.

60. Oksanen J, et al. vegan: Community Ecology Package. 2022. https://cran.r-project.org/web/packages/vegan/index.html.

